# Lack of immune homology with vaccine preventable pathogens suggests childhood immunizations do not protect against SARS-CoV-2 through adaptive cross-immunity

**DOI:** 10.1101/2020.11.13.20230862

**Authors:** Weihua Guo, Kyle O. Lee, Peter P. Lee

**Affiliations:** Department of Immuno-Oncology, Beckman Research Institute at the City of Hope, Duarte, CA, USA 91010

## Abstract

Recent epidemiological studies have investigated the potential effects of childhood immunization history on COVID-19 severity. Specifically, prior exposure to Bacillus Calmette–Guérin (BCG) vaccine, oral poliovirus vaccine (OPV), or measles vaccine have been postulated to reduce COVID-19 severity – putative mechanism is via stimulation of the innate immune system to provide broader protection against non-specific pathogens. While these epidemiological results remain inconclusive, we sought to investigate the potential role of adaptive immunity via cross-reactivity between vaccine preventable diseases (VPDs) with SARS-CoV-2. We implemented a comprehensive exploration of immune homology (including sequence homology, immune epitopes, and glycosylation patterns) between SARS-CoV-2 and all pathogens with FDA-approved vaccines. Sequence homology did not reveal significant alignments of protein sequences between SARS-CoV-2 with any VPD pathogens, including BCG-related strains. We also could not identify any shared T or B cell epitopes between SARS-CoV-2 and VPD pathogens among either experimentally validated epitopes or predicted immune epitopes. For N-glycosylation (N-glyc), while sites with the same tripeptides could be found between SARS-CoV-2 and certain VPD pathogens, their glycosylation potentials and positions were different. In summary, lack of immune homology between SARS-CoV-2 and VPD pathogens suggests that childhood immunization history (i.e., BCG vaccination or others) does not provide protection from SARS-CoV-2 through adaptive cross-immunity.

**Highlights:**

- Comprehensive exploration of immune homology for SARS-CoV-2 with 34 vaccine preventable pathogens covering all FDA-approved vaccines.
- Little to no immune homology between SARS-CoV-2 and VPD pathogens: insignificant aligned protein sequences, unmapped immune epitopes, or matched N-glycosylation sites with different glycosylation potentials and positions.
- BCG vaccination is unlikely to confer SARS-CoV-2 protection through adaptive cross-immunity.

**Graphic summary:** 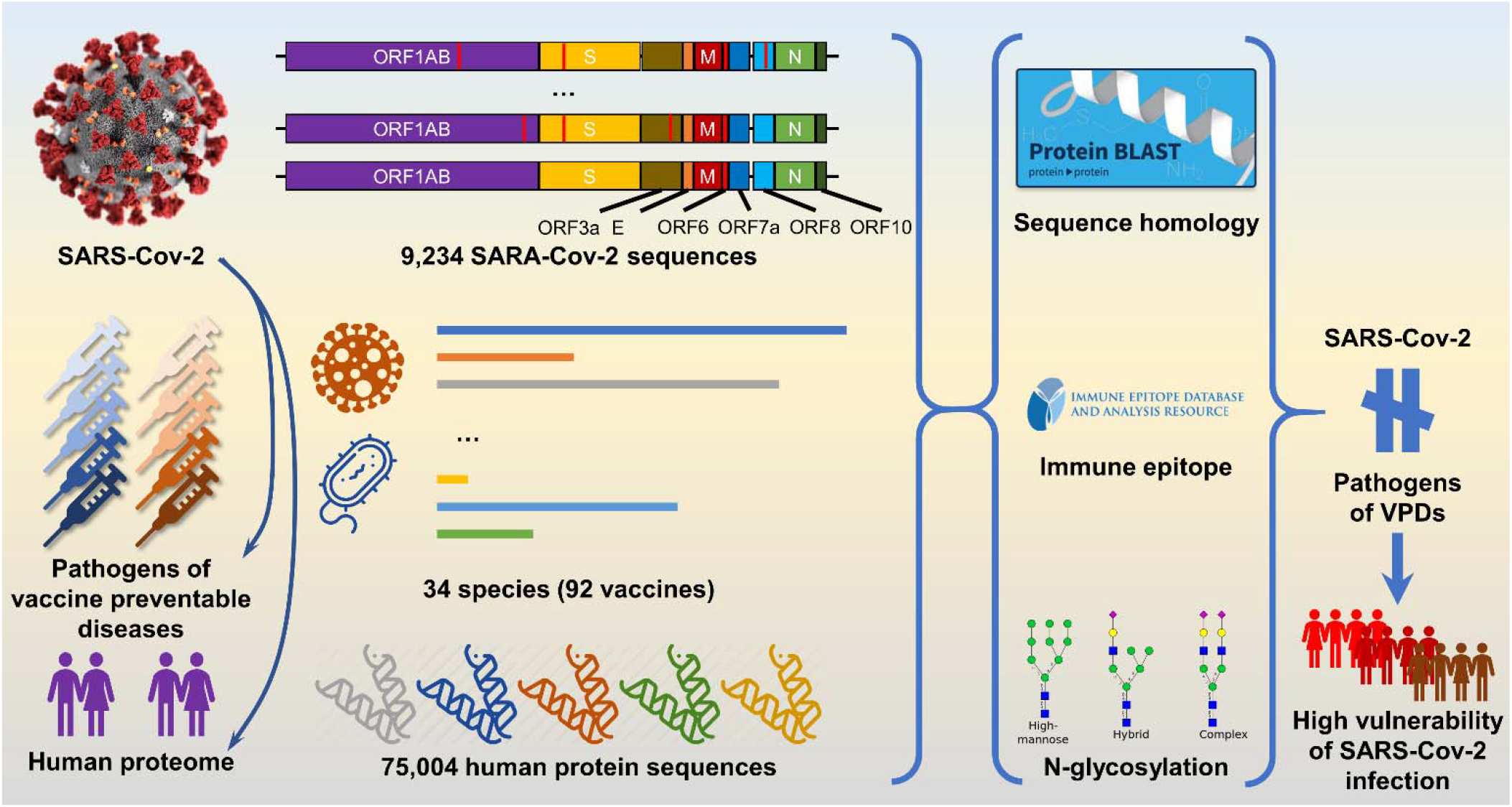

## Introduction

Severe acute respiratory syndrome coronavirus 2 (SARS-CoV-2), a novel enveloped RNA betacoronavirus, is the causative pathogen of COVID19. It has infected over 40 million people leading to more than one million deaths worldwide since December 2019 [https://covid19.who.int]. Recent epidemiological studies have investigated the potential impact of specific childhood immunization history on COVID19. Many of these focus on Bacillus Calmette–Guérin (BCG), a live attenuated vaccine for tuberculosis(Moorlag et al., 2019; Tanner et al., 2019). Several studies found that BCG vaccination history was negatively associated with COVID19 morbidity and mortality, independent of other epidemiological factors(Redelman-Sidi, 2020). Hegarty et al observed that both incidence (38.4 per million) and fatality rates (4.28 per million) of COVID19 in countries with national BCG vaccination programs are lower than in countries without (358.4 per million and 40 per million, respectively) (Hegarty et al., 2020). Miller et al. found statistically significant lower morbidities and mortalities of COVID19 in countries with universal BCG vaccination policy regardless of income (Miller et al., 2020). Interestingly, higher mortalities of COVID19 were observed in countries with a later start of universal BCG vaccination(Miller et al., 2020). Similar observations were found in several other studies using different datasets of time points and included countries or regions(Levine, 2020; Miyasaka, 2020; Ouanes et al., 2020; Shet et al., 2020). In addition, higher growth rates of COVID19 morbidity and mortality have been found in countries without BCG vaccination regardless of life expectancy and population (Berg et al., 2020; Sala et al., 2020). To mitigate effects of potentially confounding factors (e.g., stage of the COVID19 epidemic, development, rurality, population density, and age structure), Escobar et al. created a BCG index to correct for these factors and still showed significant reduction in COVID19 mortality from BCG vaccination (Escobar et al., 2020). Based on these epidemiological studies, several BCG-COVID19 related clinical trials are ongoing globally (e.g., Australia, Denmark, and United State)(Curtis et al., 2020). Results from a recent clinical trial of healthcare workers in United Arab Emirates showed that the BCG vaccinated group had no SARS-CoV-2 infection within 71 subjects, while 18/209 were infected in the un-vaccinated group(Amirlak et al., 2020). In addition to BCG vaccine, oral polio vaccine (OPV) has also been proposed to have a protective effect against SARS-CoV-2 due to prior evidence of broad protection in OPV vaccine subjects from a number of unrelated pathogens, and biological similarities between poliovirus and coronavirus(Chumakov et al., 2020). A randomized clinical trial for high-risk health care workers and first responders in New Orleans has been proposed to examine the protective effects of MMR vaccines against COVID19 (Fidel and Noverr, 2020). Potential mechanisms for this putative cross-protective effect remains unclear. While innate immunity is postulated(Netea et al., 2016, 2020), the role of adaptive immunity has not been investigated.

In contrast, other epidemiological studies showed no significant protective effects of existing vaccines to COVID19 after considering additional epidemiological factors (e.g., vaccination age, COVID19 test numbers, others). Hensel et al. found that the morbidity and mortality of COVID19 was not associated with BCG vaccination when limiting countries with high COVID19 test rates (> 2,500 tests per million inhabitants)(Hensel et al., 2020). Similar conclusions were also reported by other groups (Shivendu et al., 2020). Besides test rates of COVID19, age at vaccination and geographic regions also impact the putative protective effect of BCG vaccine. Hamiel et al compared the SARS-CoV-2 positive rates between local young adults (Israeli adults aged 35 to 41 years) with or without BCG vaccination(Hamiel et al., 2020) and found no significant difference in positive test rates. Also, no significant differences were found when comparing COVID19 mortality rates of BCG vaccinated people from Diamond Princess to the unvaccinated (Asahara, 2020). Similarly, Chaisemartin et al did not find any significant differences in numbers of COVID19 cases and hospitalized cases in Sweden when comparing the population before and after the cancelation of universal BCG vaccination policy(de Chaisemartin and de Chaisemartin, 2020). Studies also found similar growth rates of COVID19 between countries with and without universal BCG vaccination (Li et al., 2020). Masakasu found that there was no significant difference of the maximum daily increasing rate in death among countries with different BCG vaccination policies(Asahara, 2020). In addition to BCG, no significant correlations were found between COVID19 mortality and incidence of measles or rubella(Sala et al., 2020).

As these epidemiological studies are inconsistent, inconclusive, and often under powered, we approached this important hypothesis from the perspective of adaptive immunity (Redelman-Sidi, 2020). While focusing on BCG, we also broadened this analysis to all VPD pathogens with current FDA approved vaccines. We designed and implemented a comprehensive computational exploration of immune homology between SARS-CoV-2 and all VPD pathogens, focusing on three major key aspects: comparison of sequence homology, immune epitopes (including T cell and B cell epitopes), and predicted N-glycosylation (N-glyc) sites. To accomplish this, we built a proteome database for VPDs (PDB-VPD, supplementary data 1) including proteomes (n = 67) of all VPD pathogens (n = 34) based on the list of FDA-approved vaccines currently licensed for use in the United States (Table S1, Supplemental Data 1, https://www.fda.gov/vaccines-blood-biologics/vaccines/vaccines-licensed-use-united-states). To cover evolved variants of SARS-CoV-2, we compared all available protein sequences of SARS-CoV-2 from NCBI Virus database (Supplementary Data 1, n = 9,234) to the protein sequences within PDB-VPD and the human proteome (Table S2). Since BCG has been the major focus of putative cross-protective effects on COVID19, we specifically compared all available proteomes of BCG-related strains (1 wild-type strain, 9 strains for vaccination in different regions globally, Table S3, Supplemental Data 1) against all SARS-CoV-2 proteins. In addition, we also predicted all the immune epitopes for BCG-related strains (Supplemental Data 3) and investigated the homology between immune epitopes of BCG-related strains and SARS-CoV-2 proteins. To search potential pre-existing immune epitopes from vaccination for SARS-CoV-2, we extracted the human epitope sequences of both T cells (n = 51,310) and B cells (n = 10,414) for all VPD pathogens from the Immune Epitope Database(Vita et al., 2019) (Table S4) and compared these epitope sequences against SARS-CoV-2 protein sequences (Supplemental Data 4). Lastly, N-glycosylation sites of both SARS-CoV-2 protein sequences and protein sequences in PDB-VPD were predicted by NetNGlyc 1.0 server(Gupta et al., 2004). We categorized predicted N-glyc sites based on the tripeptides (aka N-glycosylation sequon). Within sites with a consensus tripeptide, we compared the glycosylation potentials and site locations within the full peptide chain between SARS-CoV-2 and VPD pathogens (Supplemental data 5).

## Results

### Sequence homology of SARS-CoV-2 compared to VPD pathogens and human

To quantitatively examine the sequence homology between SARS-CoV-2 and VPD pathogens, we included three key parameters, i.e., alignment length, identity percentage, and E-values, from BLASTP results (with default settings of NCBI blast+ package(Camacho et al., 2009)). We collected 9,234 protein sequences of SARS-CoV-2 from NCBI virus database to cover available genetic variants of SARS-CoV-2. All SARS-CoV-2 protein sequences were organized and categorized into the genome structure of the Wuhan-Hu-1 (Fig. 1A, NC_045512.2(Wu et al., 2020)). According to the taxonomy of SARS-CoV-2, we selected SARS-CoV as positive controls to further justify the relative sequence homology.

**Figure 1.**
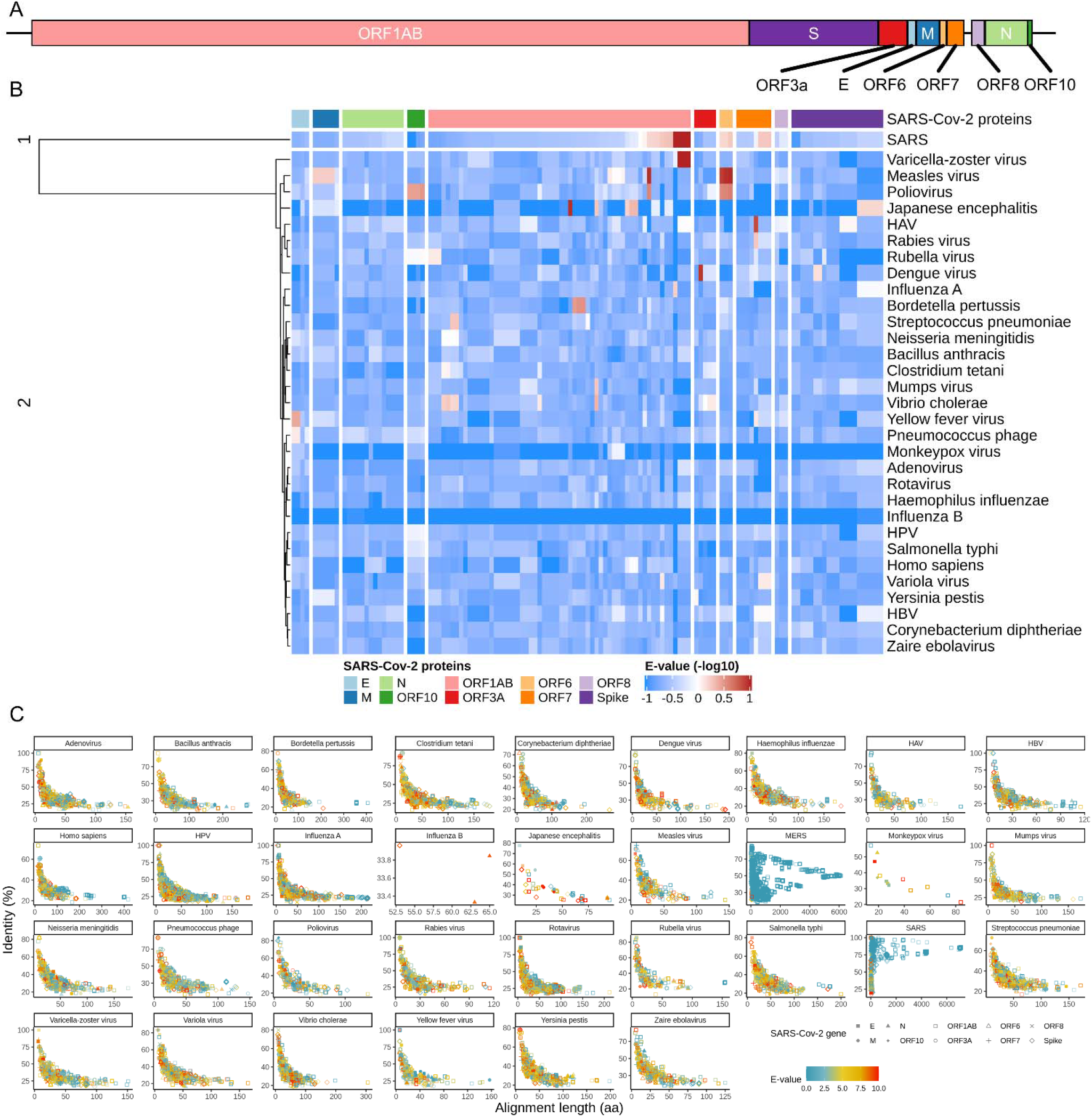
Sequencing homology between SARS-Cov-2 and pathogens of vaccine-available diseases. A) Genome structure of SARS-Cov-2 (NC_045512.2). B) Heatmap of average E-values crossing species of interests (N = 33) with hierarchical clustering. The average E-values are calculated based on each protein sequence of SARS-Cov-2 and each pathogen. E-values are log-transformed (-log10) and clustered by hierarchical clustering (details in methods). C) Correlation of E-values, identity percentages, and alignment lengths.

Consistent with previous studies(Grifoni et al., 2020), SARS-CoV has high proteome sequence homology to SARS-CoV-2 with long alignments, high identities, and low E-values (Fig. 1B & Fig. S1, supplemental data 2). All VPD pathogens have substantially lower sequence homologies to SARS-CoV-2 as compared to SARS-CoV. Average E-values between VPD pathogens and SARS-CoV-2 proteins were calculated to represent the “distances” between VPD pathogens and SARS-CoV-2 (Fig. 1B). Hierarchical clustering was implemented (Fig. 1B) and all the VPD pathogens were stratified into an independent cluster (cluster 2 in Fig. 1B) when comparing to SARS-CoV as a control, indicating the instinct differences between VPD pathogens and SARS-CoVs. We correlated three key parameters of sequence homology in Fig. 1C. A clear pattern with lower E-values (blue) and longer alignments (right) has been found in SARS-CoV, which again demonstrated the high sequence homology between SARS-CoV-2 with SARS-CoV and MERS-CoV. In contrast, VPD pathogens show elbow-like patterns (Fig. 1C) which present the negative correlations crossing alignment length, identity percentage, and E-value, indicating that no long or significant alignments were found to SARS-CoV-2 as compared to SARS-CoV or MERS-CoV. These negative homology results help explain why the unexposed population, regardless of vaccination history, is highly vulnerable to SARS-CoV-2. One key reason is the highly different proteomes between the SARS-CoV-2 and prior encountered pathogens related VPDs.

### Sequence and epitope homology of SARS-CoV-2 compared to BCG strains

BCG vaccine has been proposed to provide protective effects against COVID19 by stimulating the innate immunity(Netea et al., 2020). However, possible involvement of adaptive immunity has not been investigated. In our study, we collected all available proteomes of BCG vaccines including 1 wild-type *Mycobacterium bovis* strain and 9 vaccination strains. We also include the wild-type Mycobacterium tuberculosis and SARS-CoV proteomes as comparisons (Fig. 2A). Average E-values were calculated between BCG related strains and SARS-Cov-2 proteins, and hierarchical clustering was implemented to unbiasedly stratify the BCG related strains and SARS-Cov (Fig. 2A). Likewise, all the BCG related strains were separated from SARS-Cov as a control. Vastly lower sequence homology of SARS-Cov-2 (including shorter alignments, higher E-value, and lower identity) was observed in all the BCG-related strains (Fig. S2). Similarly, by correlating these three parameters, the same elbow-like pattern found within VPD pathogens was identified in all BCG-related strains (Fig. 2B). Such low sequence homology argues against adaptive cross-immunity via specific T or B cell memory for protective effects to SARS-CoV-2.

**Figure 2.**
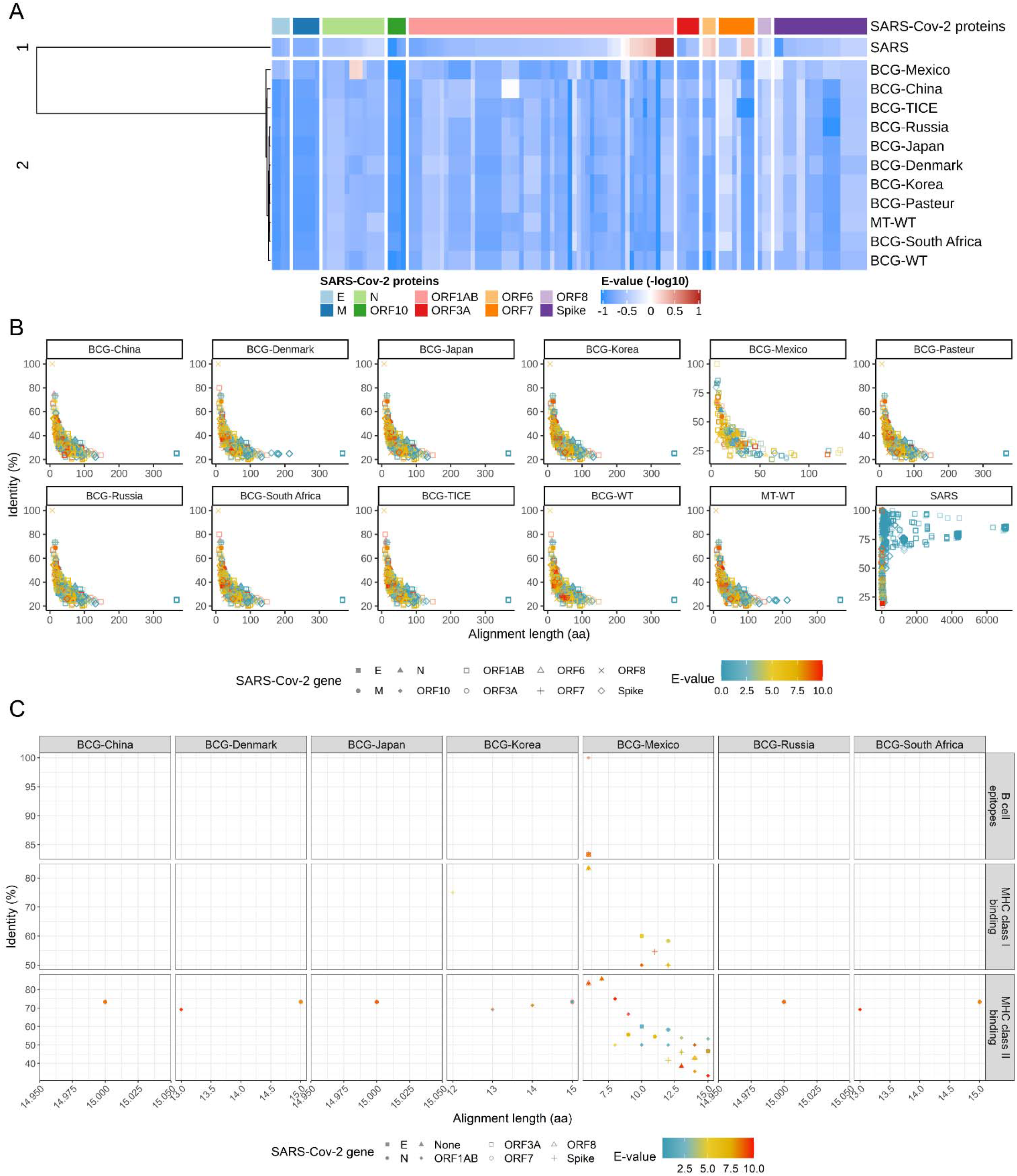
Immune homology between SARS-Cov-2 and BCG-related strains. A) Heatmap of average E-values crossing all the BCG-related strains with hierarchical clustering. The average E-values are calculated based on each protein sequence of SARS-Cov-2 and each BCG-related strain. E-values are log-transformed (-log10) and clustered by hierarchical clustering (details in methods). B) Correlation of E-values, identity percentages, and alignment lengths for each pathogens of interests. C) Homology between predicted immune epitopes of BCG related strains (including MHC class I [middle row], MHC class II [bottom row], and b cell [top row] epitopes) and SARS-Cov-2 based on sequence comparison and key parameter correlation. Total 39,331 aligned fragments were found and the aligned fragments with the same homology parameters were overlapped here. Since BCG-Pasteur and BCG-TICE have been widely applied in many countries globally, the most popular alleles cannot be selected based on countries and these two strains have been ignored in the immune epitope prediction and comparison.

Immune cells process proteins and present immune epitopes with specific amino acid sequences, which play a pivotal role in immune surveillance. To further investigate potential immune epitope homology between BCG-related strains and SARS-CoV-2, we predicted immune epitopes (including peptides binding to MHC class I [23,350,841 unique predicted peptides], peptides binding to MHC class II [6,079,665 unique predicted peptides], and B cell epitopes [7,681,061 predicted epitopes], Fig. 2C) using IEDB analytic tools (See methods, (Vita et al., 2019)) with the corresponding most common HLA alleles (top 3) for each strain (supplemental table S2). To maximize the sensitivity of this prediction, we did not filter predicted peptides (or epitopes) with any additional criteria (Supplemental data 3). Low immune homology was observed consistent with low sequence homology between BCG-related strains and SARS-CoV-2 (Fig 2C).

### Immune epitope homology of SARS-CoV-2 compared to other VPD pathogens

In addition to BCG strains, we conducted a comprehensive search of other VPD pathogens using the Immune Epitope Database (IEDB) (Vita et al., 2019). Firstly, we identified 51,310 T cell epitopes from 237 strains of VPD pathogens, and 10,414 B cell epitopes from 157 strains of VPD pathogens (Table S3). To search for shared immune epitopes with SARS-CoV-2, we aligned the 9,234 protein sequences of SARS-CoV-2 to analyze both T cell and B cell epitopes with NCBI blast+ BLASTP function. Alignment length, identity percentage, and E-value have been used to quantify similarities between potential aligned epitopes and SARS-CoV-2 (supplemental data 4). SARS-CoV served as the control group to justify relative similarities. Consistent with previous studies, SARS-CoV-2 has highly matched epitopes for both T cells and B cells to SARS-CoV (Fig. 3A), which were enriched in spike protein (light purple) and nucleoprotein (light green). Compared to SARS-CoV, VPD pathogens show far less matched epitopes for both T cells and B cells and the alignment quality of these matched epitopes are lower relatively. Since epitopes are generally shorter, we ranked the species of interested based on identity percentages. There were no significant mapped epitopes found compared to SARS-CoV, as consistent with sequence homology analysis. To further analyze the detailed sequence homology of matched epitopes, we correlated the three key parameters for T cell (Fig. 3B) and B cell (Fig. 3C) epitopes. SARS-CoV-2 has potentially shared T cell epitopes and B cell epitopes from spike protein (x dot) and nucleoproteins (filled triangle) with SARS-CoV. Compared to SARS-CoV, there were no significantly mapped epitopes found from VPD pathogens.

**Figure 3.**
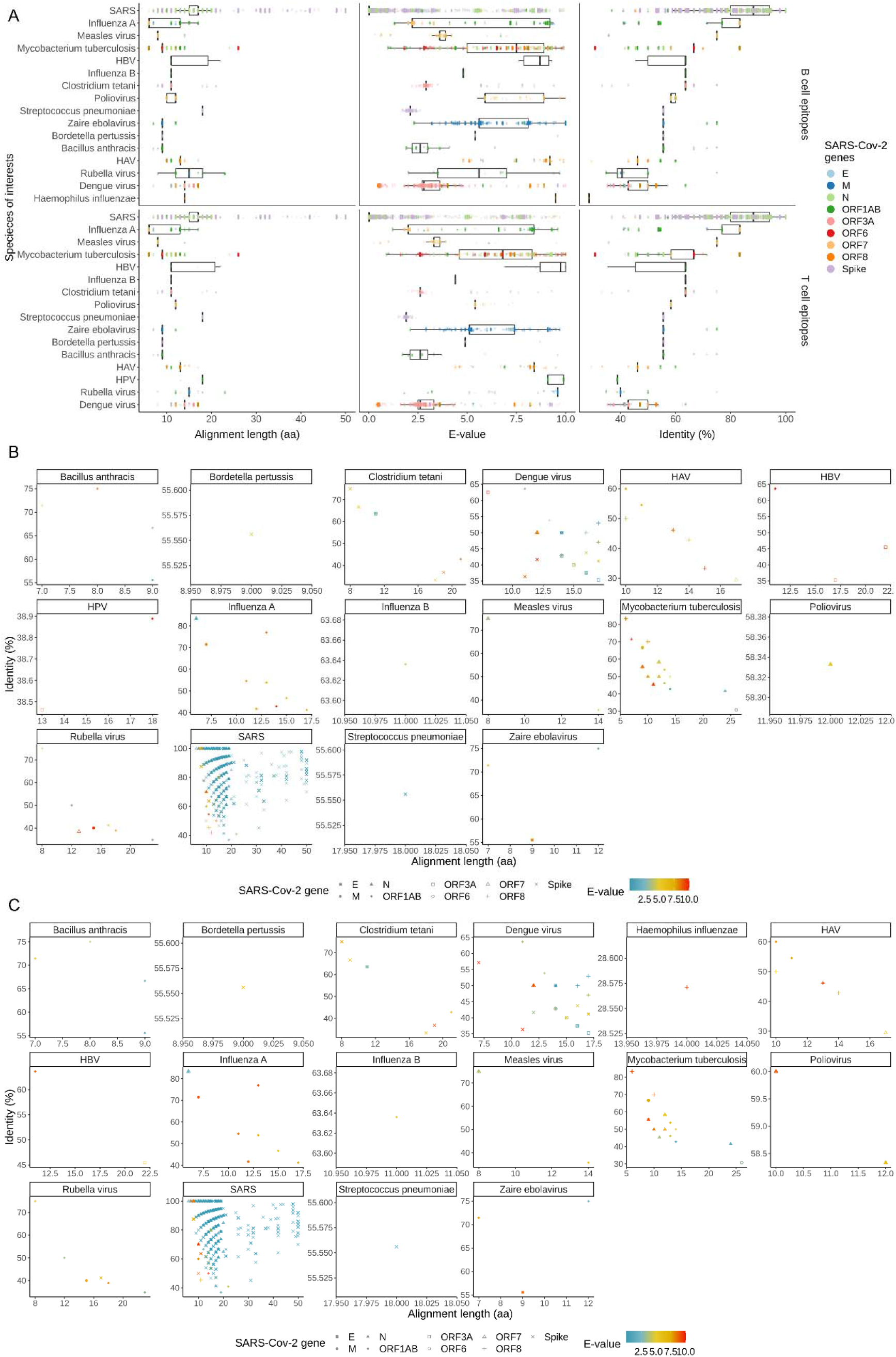
Alignments between SARS-CoV-2 and immune epitopes of vaccine-approved pathogens. A) Comparison of alignment length (left), E-value (middle), and identity percentage (right) within T cell (lower) and B cell (upper) epitopes crossing species of interests. The species order (x axis order) is sorted by the alignment length. Average is shown in the boxplot, and outliers are ignored. B) Correlation of E-values, identity percentages, and numbers of aligned AA residues for T cell epitopes. C) Correlation of E-values, identity percentages, and numbers of aligned AA residues for B cell epitopes.

### Compare predicted N-glycosylation patterns between SARS-CoV-2 and VPD pathogens

N-glycosylation is an important post-translational modification step and is associated with immune recognition and immunogenicity. To comprehensively compare the N-glycosylation patterns of SARS-CoV-2 to VPD pathogens (including SARS-CoV as control), we used NetNGlyc 1.0 server (Gupta et al., 2004) to estimate the N-glycosylation potential for all possible N-glycosylation sequons (by default settings only including Asn-Xaa-Ser/Thr sequons [i.e., tri-peptides potentially being glycosylated], where Xaa is any amino acids except proline) in 26 key proteins of SARS-CoV-2 and all proteins of VPD pathogens (supplement data 5). Firstly, we identified matched sequons from VPD pathogens which had the same tri-peptides as sequons from SARS-Cov-2 proteins. Then, we grouped the identified matched sequons from VPD pathogens based on each sequon of each SARS-CoV-2 protein. Namely, if tri-peptides of a sequon from a VPD pathogen were the same as tri-peptides of a sequon from a SARS-CoV-2 protein, we labeled this sequon from the VPD pathogen as a matched sequon to the corresponding sequon from SARS-Cov-2. In this case, each sequon of each SARS-Cov-2 protein could have one or more matched sequons from VPD pathogens with corresponding predicted N-glycosylation potentials. Then we calculated average N-glycosylation potentials for the matched sequons from VPD pathogens based on the corresponding SARS-Cov-2 sequons (Supplemental data 6). Next, sequon differences were calculated between the average potentials of VPD pathogens and corresponding potentials of SARS-CoV-2 (visualized in Fig. 4A, supplemental data 6).

**Figure 4.**
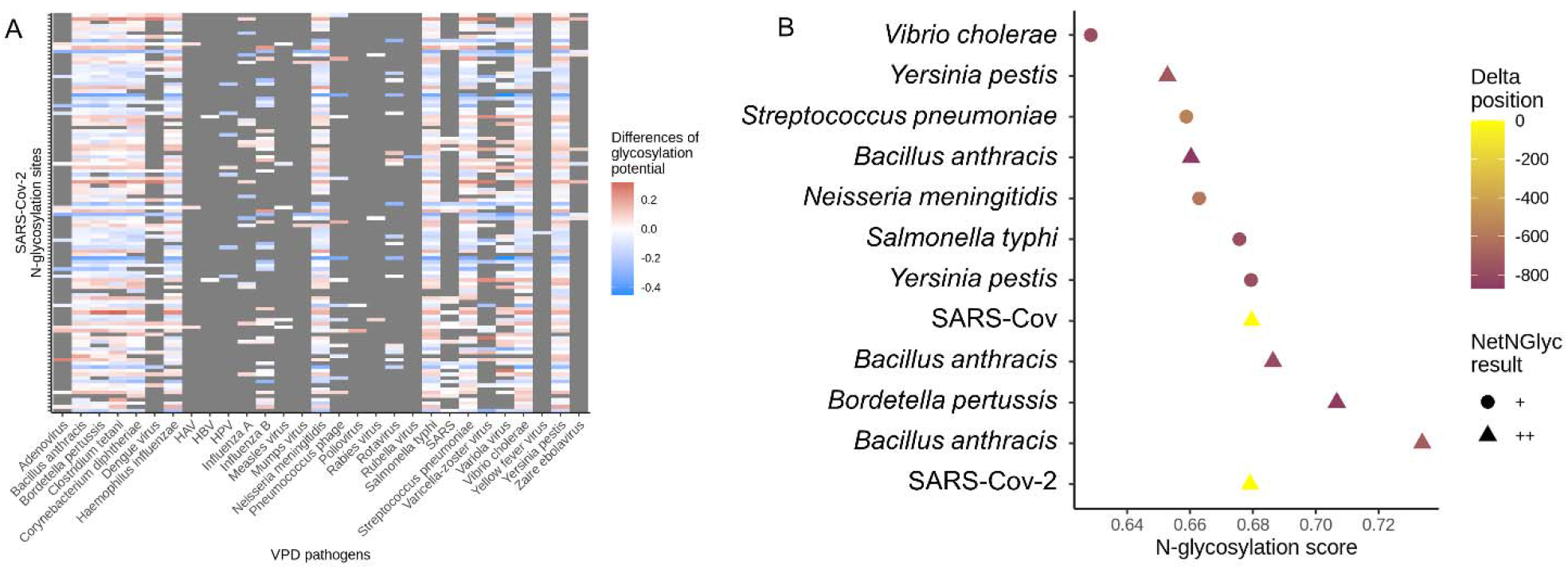
Comparison of predicted N-glycosylation patterns between SARS-CoV-2 and VPD pathogens. A) Heatmap of differences of average glycosylation potential crossing all the VPD pathogens. Each row represents a predicted N-glycosylation sequon of a SARS-CoV-2 protein (26 proteins within total 58,658 sequons, supplemental data 6). The differences of N-glycosylation potentials were calculated from the average glycosylation potentials of each pathogen (each column) and the potential of the corresponding matched sequon from each SARS-CoV-2 protein. Grey tiles represent no matched sequons from VPD pathogens (column) for that sequon from the SARS-Cov-2 protein (row). B) The matched N-glycosylation sequon (i.e., NESL in SARS-CoV-2 spike protein) with similar N-glycosylation potential (N-glycosylation score). Delta position is the differences of the amino acid positions between the NESL sequon in SARS-CoV-2 spike protein and the NESL sequon in matched proteins of VPD pathoges.

In general, by counting matched sequons from each VPD pathogen, we observed that 55.2% of VPD pathogens (N = 16, e.g., HAV, HBV, and HPV, supplemental data 6) could cover less than 30% of sequons from SARS-CoV-2 proteins. In other words, more than 70% of the total sequons from SARS-CoV-2 proteins could not be found within these 16 VPD pathogens (55.2% of the total sequons from VPD pathogens). For the matched sequons, differences of N-glycosylation potentials varied dramatically (maximum potential difference of 0.32 with standard deviation of 0.12), indicating the N-glycosylation potentials of matched sequons from VPD pathogens are generally distinguished from the potentials of SARS-CoV-2 sequons with the same tri-peptides (Fig. 4A). In addition to different N-glycosylation potentials, sequon location (i.e., amino acid position within the whole protein) also plays an important role in the N-glycosylation process. We found that the locations of matched sequons of VPD pathogens were different from locations of the corresponding SARS-CoV-2 sequon (supplemental data 5).

As an example, we extracted the comparison results of spike protein only (YP_009724390(Wu et al., 2020), Fig. 4B), since the spike protein of SARS-CoV-2 plays pivotal roles in infecting host cells. Consistent with previous studies, 22 N-glycosylation sequons were predicted by NetNGlyc in SARS-CoV-2 spike protein. Within these 22 sequons from the SARS-Cov-2 spike protein, only one predicted sequon (NESL at 1,194) had matched sequons from VPD pathogens. We correlated the N-glycosylation scores, sequon location differences, and N-Glyc results in Fig. 4B. Matched sequons from VPD pathogens (closest to x axis as the standard for other sequons from VPD pathogens) were ranked by N-glycosylation potentials and colored by absolute sequon location differences. As a control, NESL sequon from SARS-CoV had the most similar N-glycosylation pattern (i.e., similar N-glycosylation potential and similar sequon location) when compared to NESL sequon from SARS-CoV-2. Other than SARS-CoV, *Yersinia pestis* and *Bacillus anthracis* had several NESL sequons with similar N-glycosylation potentials compared to NESL sequon from the SARS-CoV-2 spike protein. However, the locations of these sequons from the corresponding proteins in VPD pathogens are different from the locations in the SARS-CoV-2 spike protein (> 600 aa). Altogether, these results demonstrated that SARS-CoV-2 has low N-glycosylation homology to VPD pathogens.

## Discussion

This study focused on computationally investigating the sequence-based immune homology between SARS-CoV-2 and all pathogens with FDA-approved vaccines to investigate potential cross-protective effects of childhood immunizations to COVID19. We built a comprehensive protein sequence database for all pathogens with FDA approved vaccines and explored sequence homology between SARS-CoV-2 and VPD pathogens using both unsupervised and supervised approaches. Only short alignments (< 50 bp) with low identity were found between SARS-CoV-2 and VPD pathogens. To further investigate the immune homology between SARS-CoV-2 and VPD pathogens, we found that no immune epitopes from VPD pathogens had high sequence homology to SARS-CoV-2 proteome. Lastly, we compared the N-glycosylation patterns between SARS-CoV-2 and VPD pathogens but only distinguished patterns were found with different N-glycosylation potentials and sequon locations. Altogether, lack of immune homology between SARS-CoV-2 and VPD pathogens suggests that childhood immunizations do not protect against SARS-CoV-2 through adaptive cross-immunity.

For this study, we built a unique, precise, and comprehensive protein sequence database for all pathogens with FDA approved vaccines. As the most comprehensive vaccine information database, VIOLIN (Vaccine Investigation and Online Information Network) also provides curated data from basic and clinical research databases (e.g. NCBI), as well as various bioinformatic tools for vaccine development(He et al., 2014; Xiang et al., 2008). However, some of the data from VIOLIN (e.g., Virmugen) are outdated and included vaccines in development. Here, we collected the latest proteome and immune epitopes for all pathogens with FDA approved vaccines. While there are additional human pathogen bioinformatic databases (e.g., ViPR (Pickett et al., 2012), PATRIC (Davis et al., 2020), dbDiarrhea (Ramana and Tamanna, 2012), etc) providing curated data related to infectious diseases, these databases do not directly provide information related to corresponding vaccines or do not include all pathogens with vaccines. Going forward, our database could be used to rapidly search for commercially available approved vaccines that may provide cross-protection against future pathogens based on immune homology. This strategy could help flatten the curve while waiting for a specific vaccine to go through clinical development.

Although other studies have investigated immune homology between SARS-CoV-2 with other pathogens, their focus had been within the betacoronavirus genus. Through computational analyses, Grifoni et al. (Grifoni et al., 2020) identified potential shared immune epitopes between SARS-CoV-2 with other related coronavirus species: 10 B cell epitope regions and 45 T cell epitope regions of SARS-CoV-2 were found to be similar to corresponding SARS-CoV epitopes. Phylogenetic network analysis has also been used to analyze SARS-CoV-2 genomes to learn the spatial evolution of documented COVID19 cases, in which SARS-CoV has been shown to be the closest species to SARS-CoV-2 based on protein sequence homology(Forster et al., 2020). In addition to computational studies, Poh et al. found a pan-SARS B-cell epitopes (i.e., IgG immunodominant regions) from SARS-CoV-2 spike glycoprotein that can be recognized by sera from COVID19 convalescent patients (Poh et al., 2020). In addition, three linear mimotopes with matched peptides in the ORF1ab protein and spike glycoprotein of SARS-CoV-2 have been found by Shivarov et al., which can also be found in other coronaviruses (i.e., SARS-CoV, HCoV 229E and OC43) (Shivarov et al., 2020). Mateus et al. showed that a set of T cell epitopes from human blood samples before SARS-CoV-2 was discovered in 2019 can be mapped to SARS-CoV-2 the genome, and more importantly, these epitopes are cross-reactive with similar affinity to SARS-CoV-2 and common cold coronaviruses (Mateus et al., 2020). Another study found that overlapping C-terminal S epitopes from peripheral blood of patients with COVID19 and SARS-CoV-2-unexposed healthy donors had a higher homology to the spike glycoprotein of human endemic coronaviruses (le Bert et al., 2020). All of these studies provide strong evidence that prior infection with coronaviruses may lead to adaptive cross-immunity.

Since our study focused on immune homology based on protein sequence alignments, our findings pertain to adaptive immunity only. Innate immunity may still be altered by certain vaccines and may confer cross-protection against SARS-CoV-2. In this regard, the BCG vaccine has been considered promising with general cross-protective effects to various respiratory pathogens. It is suggested that childhood BCG vaccination could lead to reduced morbidity and/or mortality from COVID19 many years later(Berg et al., 2020; Curtis et al., 2020; Escobar et al., 2020). The basis for this protective effect is thought to be altered innate immunity – termed “trained immunity”(Netea et al., 2016, 2020). An alternative possibility is low-level cross-protection mediated by cross-reactive T cells or antibodies. Our study addressed this possibility and found no evidence for any significant immune cross-reactivity between SARS-CoV-2 with BCG - or other VPD pathogens. If the association between BCG vaccination and lower COVID19 severity is true, our results would support innate immunity as the putative mechanism for protection. While most myeloid cells have short life-span and no memory, potential exceptions include tissue-resident macrophages (Feuerstein et al., 2020; Wendeln et al., 2018; Xing et al., 2020) and NK cells (Cerwenka and Lanier, 2016; O’Leary et al., 2006; Pahl et al., 2018; Peng and Tian, 2017). Epigenetic-mediated non-specific immune changes have been shown to last at least one year after BCG vaccination(Uthayakumar et al., 2018). While use of BCG vaccine to transiently modulate innate immunity to reduce COVID risk is currently being tested, our results suggest that currently available FDA approved vaccines are unlikely to offer cross-protective adaptive immunity to COVID19.

### STAR methods

#### Key resources table

| RESOURCE | SOURCE | IDENTIFIER |
| --- | --- | --- |
| Biological data |  |  |
| Protein sequences of SARS-CoV-2 | NCBI Virus database | https://www.ncbi.nlm.nih.gov/labs/virus/vssi/#/virus?SeqType_s=Protein&VirusLineage_ss=Severe%20acute%20respiratory%20syndrome%20coronavirus%202,%20taxid:2697049 |
| Proteomes of VPD pathogens | NCBI Virus database & Uniprot Proteome database | https://www.ncbi.nlm.nih.gov/labs/virus/vssi/#/ & https://www.uniprot.org/proteomes/ |
| Immune epitopes | Immune Epitope Database | https://www.iedb.org/database_export_v3.php |
| Software and Algorithms |  |  |
| BLASTP v2.10.0 + | (Camacho et al., 2009) | https://www.ncbi.nlm.nih.gov/guide/howto/run-blast-local/ |
| Immune epitope prediction (including MHC class I, MHC class II, and b cell) | (Vita et al., 2019) | http://tools.iedb.org/mhci/<br>http://tools.iedb.org/mhcii/<br>http://tools.iedb.org/bcell/ |
| NetNGlyc v1.0.0 | (Gupta et al., 2004) | http://www.cbs.dtu.dk/services/NetNGlyc/ |
| Python v3.7.7 | (Rossum et al., 2009) | https://www.python.org/downloads/release/python-377/ |
| R v4.0.2 | (Team, 2020) | https://anaconda.org/conda-forge/r-base |

### Method details

#### Data collection and preparation

The NCBI virus database (Brister et al., 2014) and Uniprot proteome database(Consortium, 2018) were used to collect protein sequences of pathogens related to vaccine preventable disease (including all the BCG-related strains), SARS-CoV-2 protein sequences, and the human proteome (supplemental data 1). FASTQ files of proteomes of VPD pathogens were converted to blast-ready database. T-cell and B-cell epitope sequences were downloaded from Immune Epitope Database and epitopes with human as host and from pathogens related to VPDs were semi-automatically selected for further analyses. For predicting N-glycosylation sites, all protein sequences were put into a single text file for NetNGlyc servers.

#### Investigation of immune homology for VPD pathogens

For sequence homology, each SARS-CoV-2 protein sequence was aligned to each blast-ready database by BLASTP function from NCBI blast + package with customized python scripts for automatic implementation. The alignment results (txt files, supplemental data 2) from BLASTP were concatenated into one file by python and the result visualization was implemented by customized R scripts with ggplot2 package. The e-values of each alignments were averaged based on each SARS-Cov-2 protein and each pathogen (species). The missing e-values were filled with maximum of all the available average e-values since missing e-values represented no alignments at all. Next, the negative log10-transformed average e-values were visualized in Fig. 1B and Fig. 2A and clustered with hierarchical clustering (hclust function in R programing language [embedded in ComplexHeatmap v1.10.0 package] with “complete” method and “maximum” distance options). To search similar immune epitopes, the selected T-cell and B-cell epitopes were combined to generate two FASTQ files, respectively. Similarly, these two FASTQ files have been converted to two blast-ready databases. And each SARS-CoV-2 protein sequence was aligned to both blast-ready epitope databases by BLASTP function from NCBI blast + package with customized python scripts for automatic implementation. The alignment results (txt files, supplemental data 3) from BLASTP were concatenated into one file by python and the result visualization was implemented by customized R scripts with ggplot2 package. For predicting N-glycosylation sites, customized UNIX scripts were developed to implement NetNGlyc local package for each protein sequences of interests in this work. The result of each protein sequence was stored in a specific text file (supplemental data 4). The predicted N-glycosylation sites were extracted and concatenated for SARS-CoV-2 and VPD pathogens, respectively, by customized python scripts. The comparison of N-glycosylation sites between SARS-CoV-2 and VPD pathogens was implemented by customized R scripts.

#### Prediction of immune epitopes for BCG related strains

Before predicting the peptides binding to MHC class I and MHC class II molecules, top 3 of the most frequent alleles of each locus of each BCG related strain were collected based on the major application areas of the BCG related strains (Supplemental table S3) from http://www.allelefrequencies.net/hla6006a.asp. Specifically, the “Country” option was selected based on the major application areas of the BCG related strains. The “Locus” option was screened one by one for all the loci. All the other options were kept with default settings. For each locus, the alleles were ranked by the “Allele Frequency” and the top 3 alleles were selected for prediction. To predict peptides binding MHC class I molecules, the API tools from IEDB (http://tools-cluster-interface.iedb.org/tools_api/mhci/) was used with customized scripts for each protein sequence of BCG related strains with selected most frequent alleles and four commonly used lengths (i.e., 8, 9, 10, 12). The latest recommended prediction method for MHC class I was used here with default settings(Jurtz et al., 2017)(Reynisson et al., 2020). To predict peptides binding MHC class II molecules, the API tools from IEDB (http://tools-cluster-interface.iedb.org/tools_api/mhcii/) was used with customized scripts for each protein sequence of BCG related strains with all the possible combinations of A/B chain of the most frequent HLA-DP and HLA-DQ alleles, respectively, and the most frequent HLA-DR alleles. The latest recommended prediction method for MHC class II was used here with default settings(Vita et al., 2019). To predict B cell epitopes, the API tools from IEDB (http://tools-cluster-interface.iedb.org/tools_api/bcell/) was used with customized scripts for each protein sequence of BCG related strains. The Emini prediction method for MHC class II was used here with default settings(Emini et al., 1985). To keep both high sensitivity and specificity of the predictions of immune epitopes, no additional filtering was implemented for any predicted peptides. In other words, all the predicted peptides were considered as the potential immune epitopes and compared to the SARS-CoV-2 proteome to justify the immune homology with the same sequence homology based approach described in the previous section.

## Supplemental information

**Supplemental data 1**. Proteomes used in this study (including BCG related strains).

**Supplemental data 2**. Concatenated BLASTP results for sequence homology analysis (including BCG related strains).

**Supplemental data 3**. Predicted immune epitopes of BCG related strains.

**Supplemental data 4**. BLASTP results for immune epitope search.

**Supplemental data 5**. All the predicted N-glycosylation sequons from NetNGlyc.

**Supplemental data 6**. Average N-glycosylation potentials (per VPD pathogen per SARS-Cov-2 sequon)

**Supplemental figure S1**. Comparison of alignment length (upper), E-value (middle), and identity percentage (lower) crossing species of interests (N = 33). The species order (x axis order) is sorted by the alignment length. Average is shown in the boxplot, and outliers are ignored.

**Supplemental figure S2**. Comparison of alignment length (upper), E-value (middle), and identity percentage (lower) crossing BCG-related strains (N = 10), *Mycobacterium tuberculosis*, and SARS-Cov. The species order (x axis order) is sorted by the alignment length. Average is shown in the boxplot, and outliers are ignored.

**Supplemental table S1**. Proteomes and strains with approved vaccines (excluding BCG-related strains).

**Supplemental table S2**. Proteomes of BCG related strains and the alleles used for immune epitope predictions (i.e., MHC class I and MHC class II) for each BCG related strain.

**Supplemental table S3**. Immune epitopes used in this study with corresponding strains.

## Data Availability

All the data will be uploaded as supplemental data here. The python and R scripts for this work will be available at https://github.com/weihuaguo/CoVid19_vaccine_screen with specific request before paper accepted by peer-reviewed journals.

## Acknowledges

We thank Dr. Colt A Egelston, Dr. Caroline Hoffman, and Dr. John C. Doyle for the valuable suggestions on this work.

## Author contributions

Conceptualization, P.P.L; Proteome collection, K.O.L., W.G.; Computational work, W.G.; Writing – Original draft, W.G., K.O.L.; Writing – Review & Editing, W.G., K.O.L., P.P.L.

## Declaration of interests

The authors declare no competing interests.

---

**Table S1.**
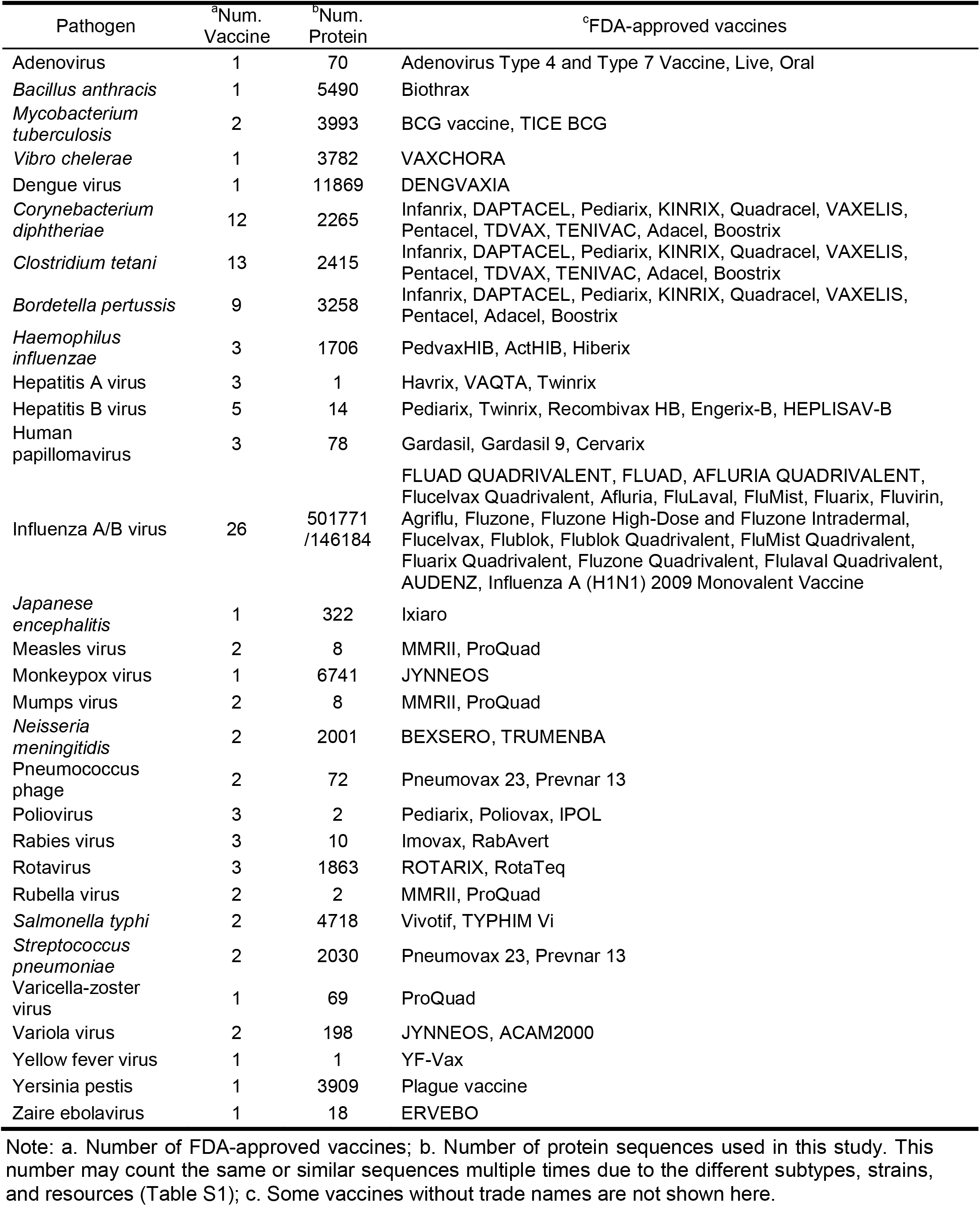
Proteomes and strains with approved vaccines.

**Supplementary figure S1.**
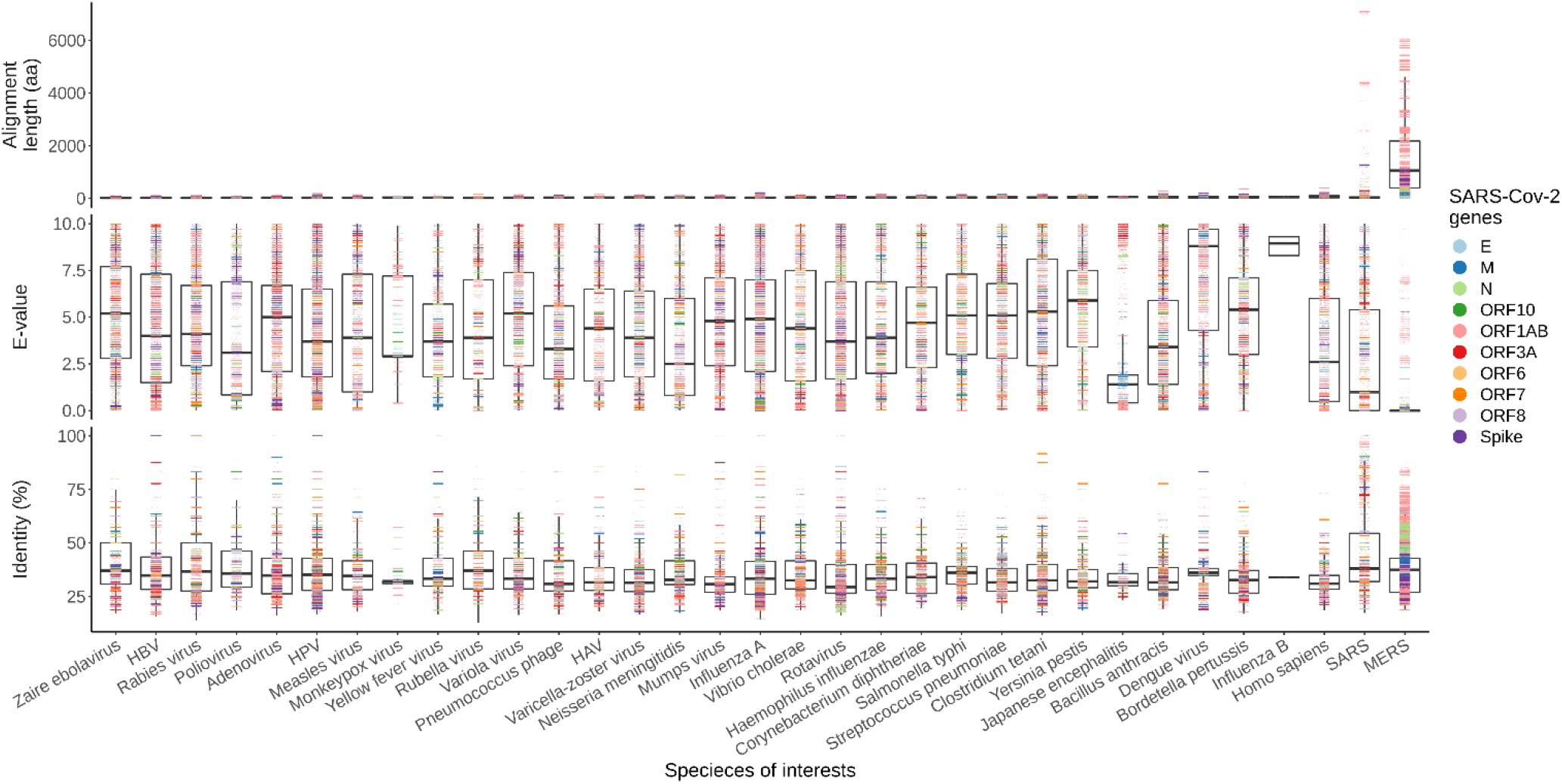
Comparison of alignment length (upper), E-value (middle), and identity percentage (lower) crossing species of interests (N = 33). The species order (x axis order) is sorted by the alignment length. Average is shown in the boxplot, and outliers are ignored.

**Supplementary figure S2.**
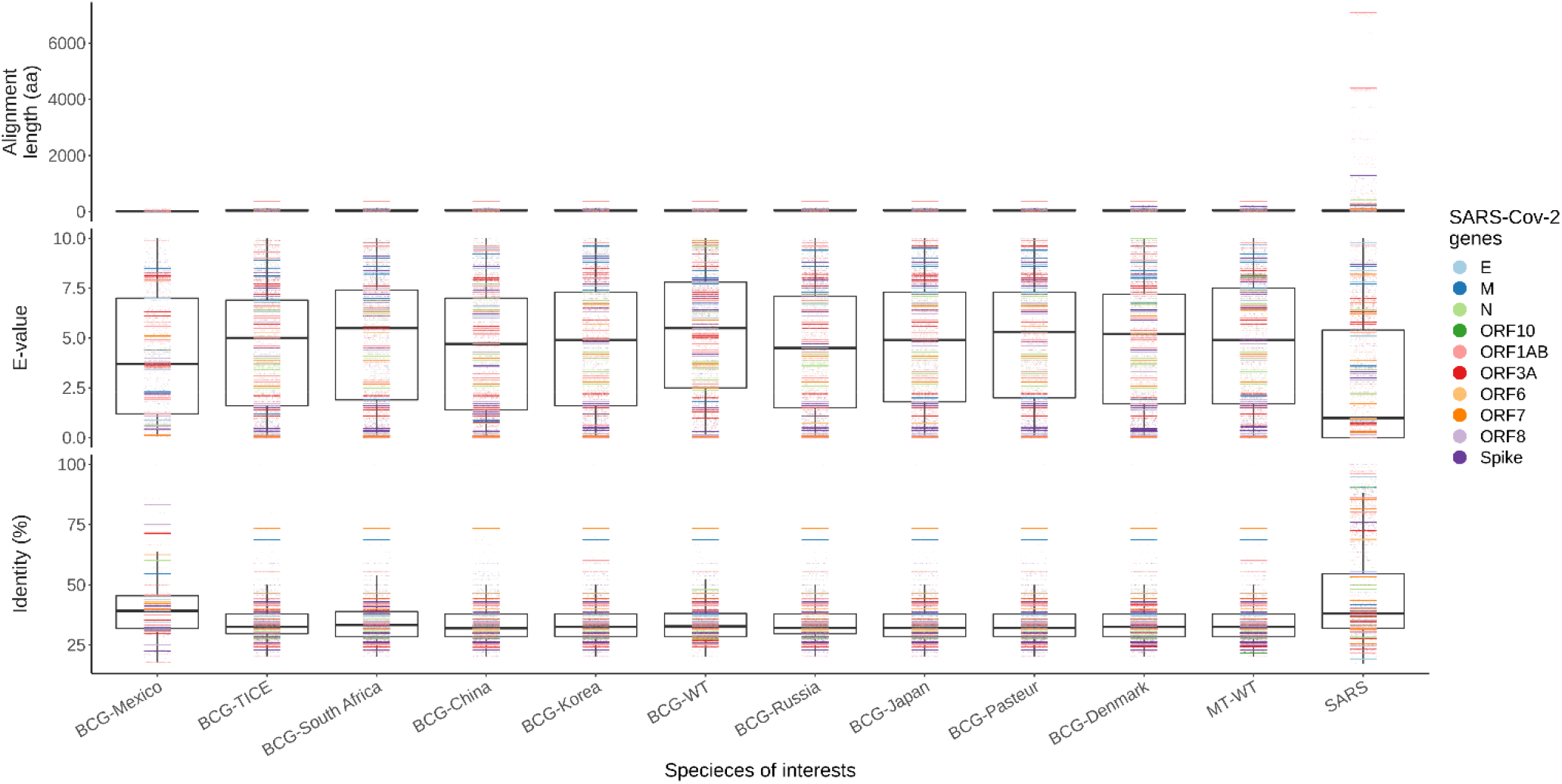
Comparison of alignment length (upper), E-value (middle), and identity percentage (lower) crossing BCG-related strains (N = 10), *Mycobacterium tuberculosis*, and SARS-Cov. The species order (x axis order) is sorted by the alignment length. Average is shown in the boxplot, and outliers are ignored.

## Notes

### Competing Interest Statement

The authors have declared no competing interest.

## References

Amirlak, I., Haddad, R., Hardy, J.D., Khaled, N.S., Chung, M.H., and Amirlak, B. (2020). Effectiveness of booster BCG vaccination in preventing COVID19 infection. MedRxiv 2020.08.10.20172288.

Asahara, M. (2020). The effect of BCG vaccination on COVID19 examined by a statistical approach: no positive results from the Diamond Princess and cross-national differences previously reported by world-wide comparisons are flawed in several ways. MedRxiv 2020.04.17.20068601.

Berg, M.K., Yu, Q., Salvador, C.E., Melani, I., and Kitayama, S. (2020). Mandated Bacillus Calmette-Guérin (BCG) vaccination predicts flattened curves for the spread of COVID19. Science Advances 6, eabc1463.

le Bert, N., Tan, A.T., Kunasegaran, K., Tham, C.Y.L., Hafezi, M., Chia, A., Chng, M.H.Y., Lin, M., Tan, N., Linster, M., et al. (2020). SARS-CoV-2-specific T cell immunity in cases of COVID19 and SARS, and uninfected controls. Nature.

Brister, J.R., Ako-adjei, D., Bao, Y., and Blinkova, O. (2014). NCBI Viral Genomes Resource. Nucleic Acids Research 43, D571–D577.

Camacho, C., Coulouris, G., Avagyan, V., Ma, N., Papadopoulos, J., Bealer, K., and Madden, T.L. (2009). BLAST+: architecture and applications. BMC Bioinformatics 10, 421.

Cernuschi, T., Malvolti, S., Nickels, E., and Friede, M. (2018). Bacillus Calmette-Guérin (BCG) vaccine: A global assessment of demand and supply balance. Vaccine 36, 498–506.

Cerwenka, A., and Lanier, L.L. (2016). Natural killer cell memory in infection, inflammation and cancer. Nature Reviews Immunology 16, 112–123.

de Chaisemartin, C., and de Chaisemartin, L. (2020). BCG vaccination in infancy does not protect against COVID-19. Evidence from a natural experiment in Sweden. MedRxiv 2020.06.22.20137802.

Chumakov, K., Benn, C.S., Aaby, P., Kottilil, S., and Gallo, R. (2020). Can existing live vaccines prevent COVID19? Science 368, 1187 LP–1188.

Consortium, T.U. (2018). UniProt: a worldwide hub of protein knowledge. Nucleic Acids Research 47, D506–D515.

Curtis, N., Sparrow, A., Ghebreyesus, T.A., and Netea, M.G. (2020). Considering BCG vaccination to reduce the impact of COVID19. The Lancet 395, 1545–1546.

Dong, E., Du, H., and Gardner, L. (2020). An interactive web-based dashboard to track COVID19 in real time. The Lancet Infectious Diseases 20, 533–534.

Escobar, L.E., Molina-Cruz, A., and Barillas-Mury, C. (2020). BCG vaccine protection from severe coronavirus disease 2019 (COVID19). Proceedings of the National Academy of Sciences 117, 17720 LP–17726.

Fidel, P.L., and Noverr, M.C. (2020). Could an Unrelated Live Attenuated Vaccine Serve as a Preventive Measure To Dampen Septic Inflammation Associated with COVID19 Infection? MBio 11, e00907–20.

Forster, P., Forster, L., Renfrew, C., and Forster, M. (2020). Phylogenetic network analysis of SARS-CoV-2 genomes. Proceedings of the National Academy of Sciences 117, 9241 LP–9243.

Grifoni, A., Sidney, J., Zhang, Y., Scheuermann, R.H., Peters, B., and Sette, A. (2020). A Sequence Homology and Bioinformatic Approach Can Predict Candidate Targets for Immune Responses to SARS-CoV-2. Cell Host & Microbe 27, 671-680.e2.

Gupta, R., Jung, E., and Brunak, S. (2004). Prediction of N-glycosylation sites in human proteins. In Preparation 0, 0.

Hamiel, U., Kozer, E., and Youngster, I. (2020). SARS-CoV-2 Rates in BCG-Vaccinated and Unvaccinated Young Adults. JAMA 323, 2340–2341.

Hegarty, P.K., Sfakianos, J.P., Giannarini, G., DiNardo, A.R., and Kamat, A.M. (2020). COVID19 and Bacillus Calmette-Guérin: What is the Link? European Urology Oncology 3, 259–261.

Hensel, J., McGrail, D.J., McAndrews, K.M., Dowlatshahi, D., LeBleu, V.S., and Kalluri, R. (2020). Exercising caution in correlating COVID19 incidence and mortality rates with BCG vaccination policies due to variable rates of SARS CoV-2 testing. MedRxiv 2020.04.08.20056051.

Korber, B., Fischer, W.M., Gnanakaran, S., Yoon, H., Theiler, J., Abfalterer, W., Hengartner, N., Giorgi, E.E., Bhattacharya, T., Foley, B., et al. (2020). Tracking Changes in SARS-CoV-2 Spike: Evidence that D614G Increases Infectivity of the COVID19 Virus. Cell.

Levine, D.I. (2020). A shred of evidence that BCG vaccine may protect against COVID19: Comparing cohorts in Spain and Italy. MedRxiv 2020.06.05.20123539.

Li, Y., Zhao, S., Zhuang, Z., Cao, P., Yang, L., and He, D. (2020). The Correlation between BCG Immunization CoVerage and the Severity of COVID19. SSRN.

Mateus, J., Grifoni, A., Tarke, A., Sidney, J., Ramirez, S.I., Dan, J.M., Burger, Z.C., Rawlings, S.A., Smith, D.M., Phillips, E., et al. (2020). Selective and cross-reactive SARS-CoV-2 T cell epitopes in unexposed humans. Science eabd3871.

Miller, A., Reandelar, M.J., Fasciglione, K., Roumenova, V., Li, Y., and Otazu, G.H. (2020). Correlation between universal BCG vaccination policy and reduced morbidity and mortality for COVID19: an epidemiological study. MedRxiv 2020.03.24.20042937.

Miyasaka, M. (2020). Is BCG vaccination causally related to reduced COVID19 mortality? EMBO Molecular Medicine 12, e12661.

Moorlag, S.J.C.F.M., Arts, R.J.W., van Crevel, R., and Netea, M.G. (2019). Non-specific effects of BCG vaccine on viral infections. Clinical Microbiology and Infection 25, 1473–1478.

Netea, M.G., Joosten, L.A.B., Latz, E., Mills, K.H.G., Natoli, G., Stunnenberg, H.G., O’Neill, L.A.J., and Xavier, R.J. (2016). Trained immunity: A program of innate immune memory in health and disease. Science 352, aaf1098.

Netea, M.G., Giamarellos-Bourboulis, E.J., Domínguez-Andrés, J., Curtis, N., van Crevel, R., van de Veerdonk, F.L., and Bonten, M. (2020). Trained Immunity: a Tool for Reducing Susceptibility to and the Severity of SARS-CoV-2 Infection. Cell 181, 969–977.

Nicola, M., Alsafi, Z., Sohrabi, C., Kerwan, A., Al-Jabir, A., Iosifidis, C., Agha, M., and Agha, R. (2020). The socio-economic implications of the coronavirus pandemic (COVID19): A review. International Journal of Surgery 78, 185–193.

O’Leary, J.G., Goodarzi, M., Drayton, D.L., and von Andrian, U.H. (2006). T cell– and B cell– independent adaptive immunity mediated by natural killer cells. Nature Immunology 7, 507–516.

Ouanes, Y., Bibi, M., Baradai, N., Boukhris, M., Chaker, K., Kacem, A., Hedhli, H., Mrad Deli, K., Sellami, A., ben Rhouma, S., et al. (2020). Does BCG protect against SARS-CoV-2 infectionlJ?: elements of proof. MedRxiv 2020.05.01.20087437.

Pahl, J.H.W., Cerwenka, A., and Ni, J. (2018). Memory-Like NK Cells: Remembering a Previous Activation by Cytokines and NK Cell Receptors. Frontiers in Immunology 9, 2796.

Peng, H., and Tian, Z. (2017). Natural Killer Cell Memory: Progress and Implications. Frontiers in Immunology 8, 1143.

Plotkin, S. (2014). History of vaccination. Proceedings of the National Academy of Sciences 111, 12283 LP–12287.

Poh, C.M., Carissimo, G., Wang, B., Amrun, S.N., Lee, C.Y.-P., Chee, R.S.-L., Fong, S.-W., Yeo, N.K.-W., Lee, W.-H., Torres-Ruesta, A., et al. (2020). Two linear epitopes on the SARS-CoV-2 spike protein that elicit neutralising antibodies in COVID19 patients. Nature Communications 11, 2806.

Redelman-Sidi, G. (2020). Could BCG be used to protect against COVID19? Nature Reviews Urology 17, 316–317.

Rossum, V., Drake, G. and, and L., F. (2009). Python 3 Reference Manual.

Sala, G., Chakraborti, R., Ota, A., and Miyakawa, T. (2020). Association of BCG vaccination policy and tuberculosis burden with incidence and mortality of COVID19. MedRxiv 2020.03.30.20048165.

Sette, A., and Crotty, S. (2020). Pre-existing immunity to SARS-CoV-2: the knowns and unknowns. Nature Reviews Immunology 20, 457–458.

Shet, A., Ray, D., Malavige, N., Santosham, M., and Bar-Zeev, N. (2020). Differential COVID19-attributable mortality and BCG vaccine use in countries. MedRxiv 2020.04.01.20049478.

Shivarov, V., Petrov, P.K., and Pashov, A.D. (2020). Potential SARS-CoV-2 Preimmune IgM Epitopes. Frontiers in Immunology 11, 932.

Shivendu, S., Chakraborty, S., Onuchowska, A., Srivastava, A., and Patidar, A. (2020). Is there evidence that BCG vaccination has non-specific protective effects for COVID 19 infections or is it an illusion created by lack of testing? MedRxiv 2020.04.18.20071142.

Tanner, R., Villarreal-Ramos, B., Vordermeier, H.M., and McShane, H. (2019). The Humoral Immune Response to BCG Vaccination. Frontiers in Immunology 10, 1317.

Team, R.C. (2020). R: A language and environment for statistical computing. R Foundation for Statistical Computing 0, 0.

Uthayakumar, D., Paris, S., Chapat, L., Freyburger, L., Poulet, H., and de Luca, K. (2018). Non-specific Effects of Vaccines Illustrated Through the BCG Example: From Observations to Demonstrations. Frontiers in Immunology 9, 2869.

Vita, R., Mahajan, S., Overton, J.A., Dhanda, S.K., Martini, S., Cantrell, J.R., Wheeler, D.K., Sette, A., and Peters, B. (2019). The Immune Epitope Database (IEDB): 2018 update. Nucleic Acids Research 47, D339–D343.

Watanabe, Y., Allen, J.D., Wrapp, D., McLellan, J.S., and Crispin, M. (2020). Site-specific glycan analysis of the SARS-CoV-2 spike. Science 369, 330 LP–333.

Wu, F., Zhao, S., Yu, B., Chen, Y.-M., Wang, W., Song, Z.-G., Hu, Y., Tao, Z.-W., Tian, J.-H., Pei, Y.- Y., et al. (2020). A new coronavirus associated with human respiratory disease in China. Nature 579, 265–269.

